# Statistical Analysis Plan for Balanced versus Saline trialists living systematic review individual patient and aggregated data meta-analysis of randomized controlled trials (BEST-Living Study)

**DOI:** 10.1101/2022.09.06.22279363

**Authors:** Fernando G Zampieri, Alexandre B Cavalcanti, Gian Luca Di Tanna, Lucas P Damiani, Naomi E Hammond, Flavia R Machado, Sharon Micallef, John Myburgh, Mahesh Ramanan, Todd W Rice, Matthew W Semler, Paul J Young, Simon Finfer

## Abstract

**Purpose:** The Balanced versus Saline Trialists (BEST) -Living Study is an ongoing living systematic review with aggregated and individual patient data meta-analysis (IPDMA) from eligible trials that assessed the effects of using balanced solutions compared with saline in critically ill adults. We herein present the search strategies for the BEST-Living Study and provide details for future analysis and presentation.

**Methods:** The report will follow the Preferred Reporting Items for Systematic Reviews and Meta-Analyses Protocols (PRISMA-P) and Preferred Reporting Items for Systematic Reviews and Meta-Analyses of Individual Participant Data (PRISMA-IPD) recommendations. Search was performed in Pubmed, EMBASE, and Cochrane Central Register of Controlled Trials (CENTRAL). The primary endpoint will be hospital mortality, which will be analyzed using a Bayesian hierarchical model. Secondary endpoints include nurvival until longest follow-up available, use of kidney replacement therapy, and intensive care unit and hospital length-of-stay. Details on the analysis plan are provided in this statistical analysis plan.

**Conclusion:** The study will provide the most up-to-date and comprehensive assessment using the best evidence available for the use of balanced solutions in critically ill patients.

**Approvals:** The undersigned and all authors have reviewed this plan and approve it as final. They find it to be consistent with the requirements of the protocol as it applies to their respective areas. They also find it to be compliant with ICH-E9 principles and, in particular, confirm that this analysis plan was final prior to any analysis of the aggregated data.

| **Name** | **Signature** | **Date** |
| --- | --- | --- |
| Fernando G Zampieri |  | 26 August 2022 |
| Prof. Simon Finfer |  | 26 August 2022 |
| Prof. Gian Luca Di Tanna |  | 26 August 2022 |

## Introduction

The Balanced versus Saline Trialists (BEST) -Living Study is an ongoing living systematic review with aggregated and individual patient data meta-analysis (IPDMA) from eligible trials that reported the effects of the using balanced solutions compared with saline in critically ill adults. The study will provide the most up-to-date and comprehensive assessment using the best evidence available for the use of balanced solutions in critically ill patients, as described in the original protocol [1]. We herein present the search strategies for the BEST-Living Study and provide details for future analysis and presentation. The report will follow the Preferred Reporting Items for Systematic Reviews and Meta-Analyses Protocols (PRISMA-P) [2] and Preferred Reporting Items for Systematic Reviews and Meta-Analyses of Individual Participant Data (PRISMA-IPD) [3] recommendations. We will use the latest PRISMA flowchart for clarity [4]. This statistical analysis plan reflects the plan for the first iteration of this living meta-analysis and may be subject to changes for future iterations.

## Search Strategies

The final search for the first iteration was performed on 1^st^ March 2022. The search strategies used for each source are shown in Table 1. Trials were considered if they included adults treated in an ICU; randomly allocated individuals or clusters of patients to the administration of balanced crystalloid solution or saline; and if study fluid (balanced or saline) was administered in the ICU for the duration of the ICU stay or for studies with a primary outcome of landmark mortality at day 28 or longer, study fluid (balanced or saline) was administered in the ICU until the time of landmark mortality. Studies that truncated use of study fluid in the ICU prior to 28 days were not included [1]. We excluded trials that used colloid as part of the intervention; trials that included surgical patients only; trials that did not report hospital mortality; trials in which loss to follow-up, excluding loss due to withdrawal of consent, exceeded 10% by hospital discharge; and trials in which the study fluids were used for fluid bolus (resuscitation) therapy only.

**Table 1.**
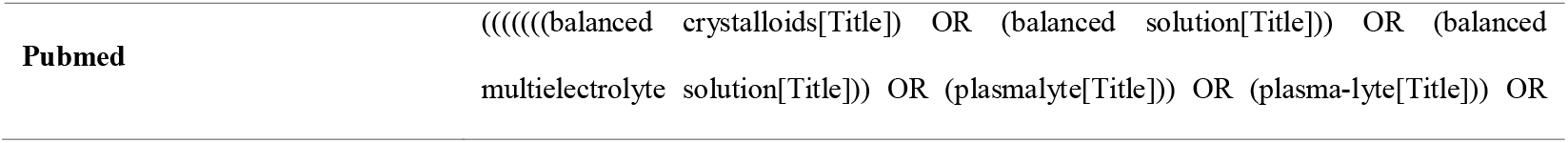
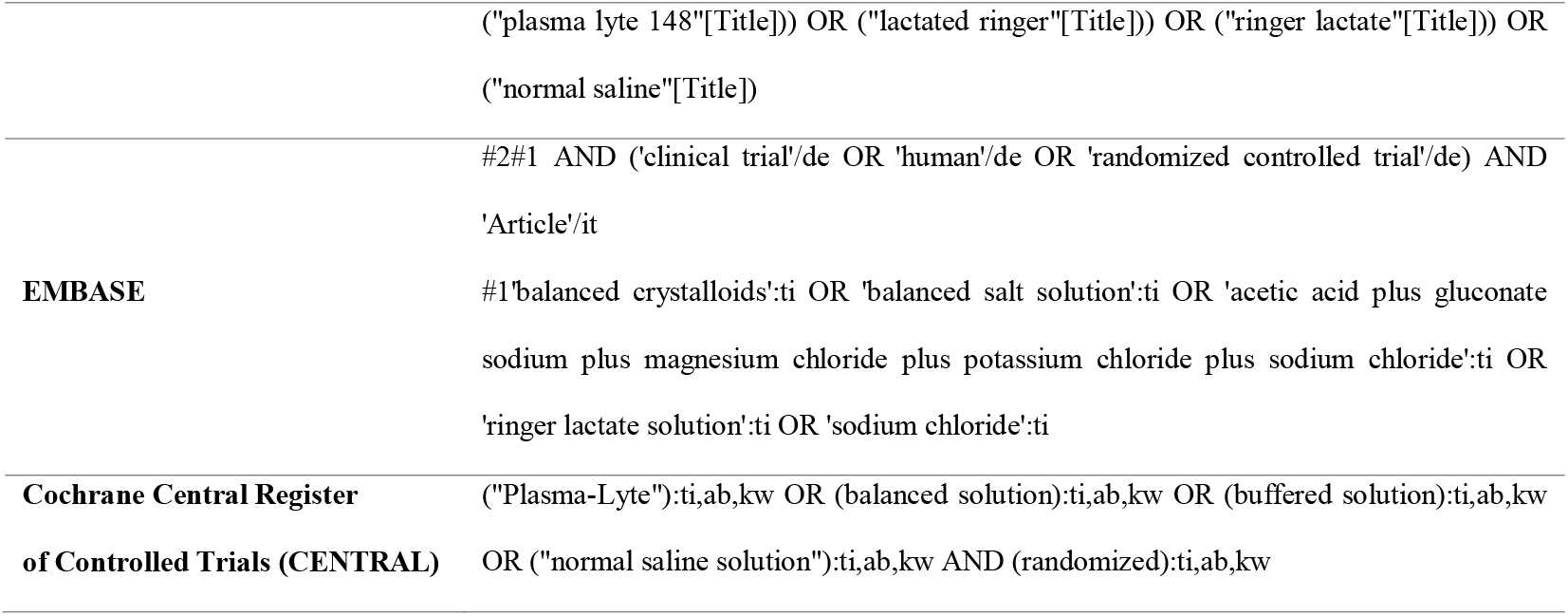
Search Strategies

Results of the searches were merged and imported into R version 4.1.3. Duplicates were removed by crosschecking information from each registry using at least two unique features of each report (for example, DOI and author name, or full text match and PMID). We also performed automated manuscript exclusions if the words “ocular”, “children”, “paediatric”, “pediatric”, “infant”, “cats”, or “dogs” appeared in the title.

## Data Sharing

The principal investigator for each eligible trial will be invited to participate and a data sharing and confidentiality agreement will be signed between the coordinating center (HCor Research Institute) and representatives of the institution responsible for each trial.

A data skeleton approved by the steering committee will be sent to the principal investigator of each trial for data population. Upon data receipt, the coordinating center will replicate the primary analysis and the main participant characteristics table of each trial (age, sex, admission type, and outcomes) to ensure data was not corrupted during transfer.

## Reporting Information of Included Trials

Baseline information and outcomes together with an overall result will be displayed according to

## Endpoints and analysis

This statistical analysis plan was developed before data from the selected trials were received and merged by the coordinating center. All analyses will be made using complete case data; that is, no imputation will be performed for missing outcomes or values from the original trials. We will perform both individual patient level meta-analysis (IPDMA) and aggregated data meta-analysis. The IPDMA performed through a one-stage approach will be the main analysis. A two-stage IPDMA will be presented as an alternative analysis for the primary endpoint (hospital mortality). We will favor the use of hierarchical modelling through the analyses; however, if convergence issues arise, we will attempt to solve them by increasing interaction depths and number of samplings. If this fails and convergence issues persists, we plan to remove the hierarchical effects for reporting and/or using the trials as a fixed-effect covariate.

### Statistical Analysis Plan for One-Stage IPDMA

#### Primary endpoint: Hospital mortality

Hospital mortality will be analyzed using a Bayesian hierarchical model with the intervention of interest (balanced solution versus saline) as a fixed-effect covariate and two layers: the ICU (or cluster) nested within trial. The hierarchical structure will be defined as different intercepts according to trial and cluster/site within trial. The main model syntax will therefore be:

~~~
main_model <-brm(hospital_mortality ∼ intervention + (1 | study/site),
family=”bernoulli”,…)
~~~

The prior for the intervention will be set as a neutral regularizing normal prior (mean 0, standard deviation of 0.355, which has 0.95 of all probability masses between an odds ratio of 0.5 to 2.0). Analyses will be performed using latest available R version [12] with packages {brms} [13] and {cmdstanr} [14]. Bayesian models will be performed using 4 chains with at least 4,000 iterations. Model convergence will be confirmed by Rhat statistics, by inspecting Markov chain plots, and by ESS values. A unique seed will be used before running the model to guarantee its reproducibility. We will use {emmeans} R package [15] to obtain the marginal odds ratio posteriors from the model. The posterior odds ratio will be summarized using the following metrics:

1. Median and 95% credible intervals (with cumulative probability plots)
2. Probability of benefit, defined as the probability that the odds ratio for hospital mortality for balanced solutions versus saline is below 1. Density and cumulative posterior probability plots will be presented.
3. Equivalence testing based on region of practical equivalence [16]. We will define equivalence region as the one encompassing a Cohen’
ss d difference from -0.025 to 0.025, which is a quarter of what could be considered a small effect size (which is usually defined as a Cohen’s d of 0.10) [16,17]. This will be done because due to the ubiquity of fluid therapy, even very small effect sizes would be considered relevant. Cohen’s d was transformed to log(OR) scale by multiplying the value by, 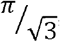 [18] resulting in a margin of equivalence for the odds ratio between 0.955-1.046; we present the percentage of the full posterior that is contained within the margins of equivalence (ROPE); values below 0.025 (2.5%) will be considered strong evidence for lack of equivalence.
4. Other effect size summaries, including risk ratio and absolute risk reductions may be obtained by sampling the model posteriors and presented.

### Sensitivity analyses for the primary endpoint

1. Multivariable/adjusted model: The model presented above will include age, sex, and admission type (surgical versus non-surgical), and sepsis. The same prior and reporting will be used.
2. Frequentist analysis: A frequentist mixed model analysis will be performed using {lme4} R package [19]. The model syntax will be the same as the primary analysis, and the results will be presented by marginal odds ratio with their 95% confidence intervals from {emmeans} package.
3. Alternative priors: Different priors will be considered, including both optimistic and pessimistic priors [20]. These priors will be designed assuming that the optimistic prior will be centered as an OR of 0.90 and the pessimistic prior will be centered at an odds of 1/0.90 (corresponding to a log(OR) of -0.105 and 0.105; the standard deviation will be set as 0.10 for both priors so that 0.15 probability of harm (for the optimistic prior) and 0.15 probability of benefit (for the pessimistic prior) are considered. We will report only median effect size with 95% credible interval and probability of benefit.

#### Key secondary endpoint: Survival until longest follow-up available

The survival analysis will be made using time-to-event models following Weibull distribution or semi-parametric models (Cox regression), under a Bayesian framework, whichever fits the data better based on posterior predictive checks. The analysis will consider hierarchical effects of site nested to trial. The results will be reported as hazard ratio and 95% credible intervals, together with the probability of direction for benefit. We will use non-informative priors for the intervention for this analysis (N(0,4)).

### Other secondary endpoints

#### Proportion of patients newly treated with kidney replacement therapy

This analysis will be performed using a model identical to the primary analysis for hospital mortality. We will only present median effect size with 95% credible intervals and probability of benefit for this analysis.

#### Length-of-stay in the ICU and Length-of-stay in the Hospital

Length-of-stay (LOS) will be analyzed using a Bayesian model using {brms}. Our intention is to use a hurdle model for this assessment. This type of model allows prediction of a binary outcome (in this case, hospital outcome as dead/alive) and a count outcome conditioned to the first prediction. This will be made by the family = hurdle_negbinomial() or hurdle_gamma() in {brms} (decision will be made according to the family that results in better posterior predictive checks); therefore, LOS will be modeled as a hurdle model with an mixture logistic part for hospital mortality and a continuous model for LOS. One model for ICU LOS and another for hospital LOS will be fit. The results will be presented as median difference with 95%, probability of benefit (any decrease in LOS), and probability of intervention being associated with a decrease in LOS of at least 1, 2, and 3 days; the model will also be adjusted for site effects. In case hurdle models do not converge, we will attempt to use days alive and free of the ICU and days alive and free of hospital truncated at 28-days as secondary analysis under a Bayesian framework using cumulative logistic models, with patients who die been given a score of - 1; in last case if model convergence issues persist, a linear mixed model will be used.

### Exploratory endpoints

The following exploratory endpoints for possible mechanistic pathways include changes in serum creatinine, sodium, chloride, and bicarbonate concentrations, and pH, over time. These results will be presented graphically as boxplots, with the x-axis representing values from baseline to days 1-7 after enrollment. We expect several trials to have data available at different timepoints (for example, one trial may have daily data up to 7 days, another during ICU discharge, and some may have no data at all). We plan to analyze these data as a Bayesian mixed model adjusted for time and trial. We will use a neutral prior with mean zero and standard deviation of 5. For simplicity, we will not consider random effects for site or trial in this analysis.

### Subgroup analyses

We will compare the effects of the intervention on hospital mortality and treatment with kidney replacement therapy in predefined subgroups. These analyses will follow the same model as the primary outcome main analysis and will be performed by splitting the sample size according to the following subgroups, already discussed in [1]. For these analyses we will only provide median and 95% credible intervals and probability of benefit, defined as the probability that odds ratio for mortality for balanced solutions compared to saline is below:

1. Patients admitted to the ICU with sepsis, as defined by each trial, compared to those without sepsis.We hypothesize that effect of balanced solutions in reducing mortality will be greater in patients with sepsis than those without sepsis.
2. Patients admitted to the ICU with traumatic brain injury (TBI) compared to those without TBI. We hypothesize that effect of balanced solutions in increasing mortality will be greater in patients with TBI than those without TBI.
3. Patients admitted to the ICU with traumatic brain injury (TBI) versus those with a non-traumatic acute brain injury versus those with no acute brain injury. We hypothesize that effect of balanced solutions in increasing mortality will be greater in patients with TBI and non-traumatic acute brain injury than in those with no acute brain injury.
4. Patients categorized by baseline plasma chloride concentration stratified in three groups (low, below 100 mmol/L; normal, 100 to 110 mmol/L; and high, above 110 mmol/L). We hypothesize that effect of balanced solutions in reducing mortality will be unaffected by baseline plasma chloride concentration.
5. Patients categorized by baseline arterial pH, in 4 groups (severe acidemia – pH < 7.20, mild acidemia – pH 7.20-7.34, normal – pH 7.35-7.45, alkalemia – pH > 7.45). We hypothesize that effect of balanced solutions in reducing mortality will be unaffected by baseline arterial pH.
6. Patients categorized by volume of saline received before enrollment, stratified into three groups (no saline received, 1 - 999 mL of saline received, or 1,000 mL or more saline received). We hypothesize that effect of balanced solutions in reducing mortality will be unaffected by receipt of saline prior to enrolment.
7. Treatment effect in women versus men using sex as reported in each trial. We hypothesize that the effect of balanced solutions versus saline will be unaffected by sex.
8. Patients recruited to cluster randomized versus individual randomization trials. We hypothesize that the effect of balanced solutions versus saline will be unaffected by unit of randomization.

### Statistical Analysis Plan for Two-Stage IPDMA

We will also perform a two-stage IPDMA in addition to the main IPDMA for the primary endpoint. Effect sizes obtained from each trial will be aggregated using a Bayesian meta-analysis for the effect size. This analysis will be performed using the {bayesmeta} package to fit normal-normal hierarchical models with a direct access to quasi-analytical posterior distributions [21].

If individual data is available for all trials, we will first estimate the effect size at each trial and then perform the Bayesian meta-analysis using uninformative priors for the mean effect size and the between trial heterogeneity [21]. If individual data for any trial is missing, we will pool the reported effect sizes to create a meta-analytic, informative prior for the mean effect size. The two-stage IPDMA will, therefore, corroborate with the primary analysis and allow the addition of information for trials whose individual data cannot be obtained.

### Statistical Analysis Plan for the Aggregated Meta-analysis

The aggregated meta-analysis will be performed for both hospital mortality and treatment with kidney replacement therapy. We will use {bayesmeta} R package [21]. We will use only neutral priors for the log(OR) of the intervention in this analysis (Normal, mean = 0, standard deviation 0.355). We will use a non-informative prior for the heterogeneity between studies estimate (tau) [21]. Median odds ratio, 95% credible interval and probability of benefit will be reported.

## Data Presentation

The results will be presented using figures and tables. We anticipate the following figures:

1. Figure 1: PRISMA flowchart
2. Figure 2: Two panels. (A) Posterior of the effect size, in log(OR) scale for the effect of the intervention on mortality; (B) Survival curves up to longest follow up according to intervention.
3. Figure 3: Posterior of the effect size for predefined subgroups, in a format that resembles a forest plot but with posteriors distributions
4. Supplementary Figures:

a. Report of the Grading of Recommendation Assessment, Development and Evaluation approach to assess overall evidence quality from included studies [22].
b. Bias assessment with a funnel plot for any analysis which includes 10 or more trials.
c. Plots for values of sodium, potassium, pH, base excess, etc. according to treatment assignment.
d. Posterior predictive checks for the models.

The following Tables will be presented

1. Table 1: Features of included patients in each trial (Table 2 in this SAP).

**Table 2.** Display of each trial baseline information and outcomes, as well as overall values
2. Table 2: Subgroup results for the primary analysis
3. Table 3: Key secondary analysis results.
4. Supplementary Tables: Results for alternative priors and other exploratory analysis

|  | Trial 1 |  | Trial 2 |  | Trial N... |  | Overall |  |
| --- | --- | --- | --- | --- | --- | --- | --- | --- |
|  | Balanced Solution | Saline | Balanced Solution | Saline | Balanced Solution | Saline | Balanced Solution | Saline |
| **BASELINE** |  |  |  |  |  |  |  |  |
| Number of patients | - | - | - | - | - | - | - | - |
| Age, mean (SD) | - | - | - | - | - | - | - | - |
| Admission Type, n (%) | - | - | - | - | - | - | - | - |
| Planned | - | - | - | - | - | - | - | - |
| Unplanned, with sepsis | -- | -- | -- | -- | -- | -- | -- | -- |
| Unplanned, without sepsis | - | - | - | - | - | - | - | - |
| Days from ICU admission to enrollment, median (IQR) | - | - | - | - | - | - | - | - |
| Sepsis, n (%) | - | - | - | - | - | - | - | - |
| Traumatic Brain Injury, n (%) | - | - | - | - | - | - | - | - |
| Other acute neurologic condition, n (%) | - | - | - | - | - | - | - | - |
| Vasopressor use, n (%) | - | - | - | - | - | - | - | - |
| Vasopressor dose, mcg/kg/min, median (IQR) | - | - | - | - | - | - | - | - |
| Inotrope use, n(%) | - | - | - | - | - | - | - | - |
| Fluid use before admission, n (%) | - | - | - | - | - | - | - | - |
| Volume of saline, mL median (IQR) | - | - | - | - | - | - | - | - |
| Volume of saline, mL median (IQR) | - | - | - | - | - | - | - | - |
| KRT at enrollment, n(%) | - | - | - | - | - | - | - | - |
| Serum Sodium, mEq/L, mean (SD) | - | - | - | - | - | - | - | - |
| Serum Chloride, mEq/L, mean (SD) | - | - | - | - | - | - | - | - |
| pH, median (IQR) | - | - | - | - | - | - | - | - |
| Base Excess, median (IQR) | - | - | - | - | - | - | - | - |
| **OUTCOMES** |  |  |  |  |  |  |  |  |
| Days from enrollment to ICU discharge, median (IQR) | - | - | - | - | - | - | - | - |
| Days from enrollment to hospital discharge, median (IQR) | - | - | - | - | - | - | - | - |
| Need for KRT, n (%) | - | - | - | - | - | - | - | - |
| Duration of KRT therapy, days, median (IQR) | - | - | - | - | - | - | - | - |
| Duration of mechanical ventilation, days, median (IQR) | - | - | - | - | - | - | - | - |
| Hospital Mortality | - | - | - | - | - | - | - | - |

## Data Availability

All data produced in the present study are available upon reasonable request to the authors

